# “There’s No Place Like Home for The Holidays:” Travel and SARS-CoV-2 Test Positivity Following Thanksgiving Weekend

**DOI:** 10.1101/2020.12.22.20248719

**Authors:** Shruti H. Mehta, Steven J. Clipman, Amy Wesolowski, Sunil S. Solomon

## Abstract

In the US, public health officials discouraged travel and social gatherings for Thanksgiving. Data suggests that many individuals did travel over the holidays, albeit in smaller numbers than previous years. Using an online panel survey of individuals across ten US states, we found that many individuals reported spending Thanksgiving outside of their home (25.9%) or at home with at least one non-household member (27.3%). Among those who were tested, those who had Thanksgiving outside their home were significantly more likely to self-report a positive PCR test for SARS-CoV-2 infection in the prior two weeks compared to those who had Thanksgiving at home with non-household members or with household members only (41.7% vs. 21.4% and 13.8%, respectively; p<0.001). Persons who had Thanksgiving outside their home and tested positive for SARS-CoV-2 participated in a median 35 (IQR: 21 - 53). non-essential activities compared to those who had Thanksgiving at home and tested positive (median 3 activities, IQR 0-13). Notably, planned travel over the December holidays was most common among those who tested positive for SARS-CoV-2 in the prior 2 weeks (66.5%) compared with 25.4% of those who tested negative in the prior 2 weeks and 11.0% among those who were not tested. While public health authorities should continue promoting messages to dissuade travel and social gatherings over the holidays, as supported by these data, it is equally important to promote messaging on how to get together in a “low-risk” manner for those who travel and plan gatherings. In particular, it is critical that those who do travel or visit with others outside their household do so cautiously and avoid or significantly minimize all other activities where they may potentially acquire and transmit infection in the weeks prior to and after their visit.

---

As the COVID-19 pandemic surged across the US in November, public health officials strongly cautioned, and indeed recommended, against travel and social gatherings for Thanksgiving[1]. Data from TSA, however, suggested that travel during the week of Thanksgiving was the highest since the beginning of the pandemic, representing only a 40% reduction from 2019 Thanksgiving travel[2]. Vehicle travel also peaked during the holiday week[3,4]. While there was substantial escalation of cases in places like California and Texas, the general consensus based on trajectories of cases counts, hospitalizations, and mortality following the holiday has been that there was no immediate nationwide surge.[3] In order to understand the full impact of the holiday, however, empirical individual-level data linking behaviors with SARS-CoV-2 positivity are needed – which to date have been lacking.

Using an online panel survey (www.dynata.com), we surveyed 7,905 individuals from 10 US states: California, Florida, Illinois, Maryland, Massachusetts, Nebraska, North Dakota, South Dakota, Texas and Wisconsin with sample sizes proportional to population size of the state ranging from 234 in the Dakotas to 1,490 in Texas. Individuals were surveyed from December 4 - December 18, 2020 representing a range of 8 - 22 days after Thanksgiving. Participants provided informed consent, were 18 years of age or older, and a resident of the state they were sampled from. Sample quotas were established based on state-level distributions of age, gender, race and income. The survey captured information on demographics, frequency of recent visits (within the past two weeks) to places of interest (POI), social distancing and mask use at these POIs, experiences with COVID-19 symptoms and SARS-CoV-2 testing in the past two weeks as well as activities for Thanksgiving and travel plans over the December holidays. Data on visits to POIs were used to calculate an activity score that summarized the number of times an individual participated in non-essential activities in the prior two weeks, this included visits to bars, restaurants, church, salons, theatres and stadiums, gyms, participation in group outdoor fitness activities and gatherings with friends and family. We compared outcomes across three groups: those who celebrated Thanksgiving only with their immediate household, those who celebrated Thanksgiving in their home but with ≥ 1 person from outside their household, and those who had Thanksgiving outside of their home.

The median age of the sample was 45 years (interquartile range [IQR]: 32 - 63); about half (49.5%) were female (Table). The majority (53.6%) were White, 13.2% were Black/African American; 24.9% self-reported Hispanic/Latino ethnicity (Table). A little over a third (38.3%) were working outside the home at the time of the survey.

Overall, 2,055 (26.0%) of 7,905 reported having Thanksgiving outside of their home with minimal variability by state (22.4% in Maryland to 30.8% in the Dakotas) and most (90.5%) reported traveling by car with 3.7% traveling by plane. An additional 2,063 (26.1%) had Thanksgiving at home but with at least one person from outside their household. The median Thanksgiving dinner size for those who had Thanksgiving outside of their home was 5 (IQR: 3 – 8) compared to 4 (IQR: 3 – 6) who had Thanksgiving at home with ≥1 non-household member and 1 (IQR: 1 – 3) for those who had Thanksgiving with their household only (p<0.001). Compared to those who had Thanksgiving with household members only, those who had Thanksgiving with non-household members or Thanksgiving outside their home were significantly more likely to report COVID-19 symptoms, testing for SARS-CoV-2, and testing positive for SARS-CoV-2 infection all in the prior 2 weeks, with the highest levels in those who had Thanksgiving outside of their home (Figure). Among those tested in the prior two weeks, those who had Thanksgiving outside their home were significantly more likely to self-report a positive SARS-CoV-2 result compared to those who had Thanksgiving with non-household members in their home or with their household only (41.7% vs. 21.4% and 13.8%, respectively; p<0.001).

Further, persons who had Thanksgiving outside their home and tested positive for SARS-CoV-2, participated in a median 35 non-essential activities in the prior 2 weeks (IQR: 21 – 53). By comparison, those who had Thanksgiving outside their home, but did not test positive participated in a median 4 activities (IQR: 2 – 8) and those who had Thanksgiving at home but tested positive for SARS-CoV-2 in the prior 2 weeks participated in a median 3 activities (IQR: 0 – 13).

In terms of plans for December, 1,514 (19.2%) were planning to either travel for the holidays or host persons from outside their household. Those who had Thanksgiving outside their home reported planning to spend the December holidays with significantly more people (median: 4; IQR: 2 – 7) than those who had Thanksgiving at their home (median: 2; IQR: 1 – 5) (p < 0.001). Notably, planned travel over the December holidays was most common among those who tested positive for SARS-CoV-2 in the prior 2 weeks (66.5%) compared with 25.4% of those who tested negative in the prior 2 weeks and 11.0% among those who were not tested.

While the majority sampled across these 10 states did not travel over Thanksgiving, it is concerning that the group that did have Thanksgiving outside of their home had a test positivity rate higher than that reported in any state in the US currently. These individuals also tended to be more active in the weeks immediately following Thanksgiving with frequent visits to bars, restaurants and gyms and are more likely to report planned gatherings in December. While we cannot definitively say whether infection was a result of Thanksgiving travel or participation in multiple recreational activities, it is clear that this small group of individuals could potentially act as “super spreaders.”[5] Moreover, it is plausible that delayed surges (as a result of population mixing post-Thanksgiving) will be observed in some states, surges that will coincide with an already expected surge from December holiday travel. Concurrently, it is important to note that among those who had Thanksgiving outside of their home, lower frequency of other non-essential activities was associated with a higher likelihood of testing negative.

Several limitations must be acknowledged. Specifically, these data cannot be considered representative of the states; however, our primary purpose was to examine associations between behaviors and outcomes for which a representative sample is not required. Further, all data is self-reported and subject to bias; however, we sampled participants in the weeks immediately following Thanksgiving and queried behaviors in the prior two weeks to minimize recall bias. All participants were not screened for SARS-CoV-2 and so, it is likely we missed undiagnosed infections; however, we observed a similar association with COVID-19 symptoms that was asked of all participants.

The holidays have traditionally been a time to get together with families. While public health authorities should continue promoting messages to dissuade travel and social gatherings over the holidays and subsequent long weekends as supported by these data, it is equally important to promote messaging on how to get together in a “low-risk” manner for those who are planning gatherings. In particular, it is critical that those who do visit with others outside their household do so cautiously and avoid or significantly minimize all other activities where they may acquire and transmit infection in the weeks prior to and after their visit.

**Figure.**
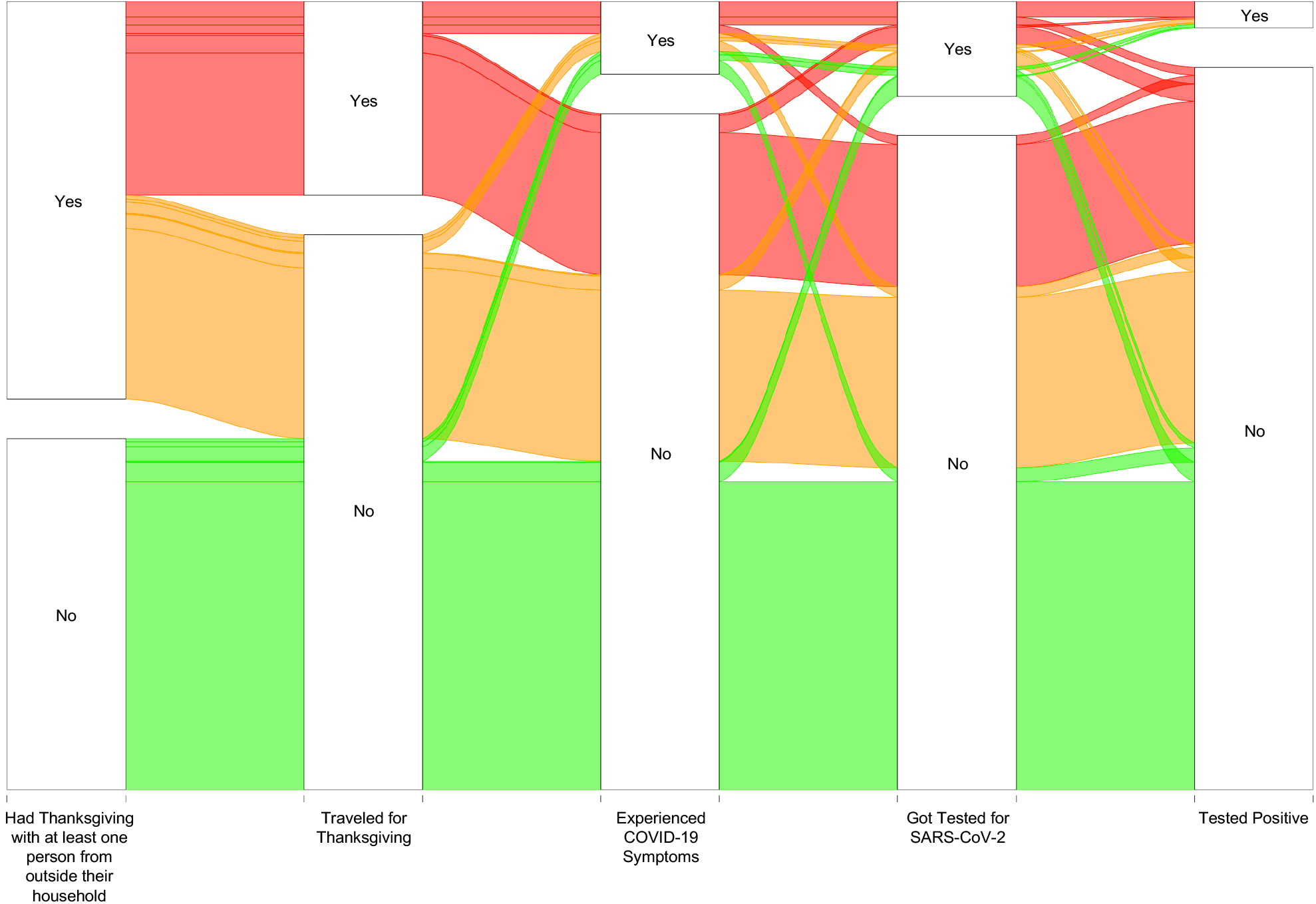
Sankey diagram showing COVID-19 symptoms, SARS-CoV-2 PCR testing, and self-reported positivity in the prior 2 weeks by Thanksgiving behavior. Participant responses are depicted in each rectangular node with flows proportional to how many individuals report that pattern of behavior. Green flows represent respondents who did not travel or have Thanksgiving with anyone outside their immediate household; orange flows represent those who had Thanksgiving in their home with at ≥ 1 person from outside their household; red flows represent those who had Thanksgiving outside their home.

**Table.**

|  | Thanksgiving<br>at Home<br>with Household<br>Members Only | Thanksgiving<br>at Home<br>with ≥ 1<br>Non-Household<br>Member | Traveled<br>Outside the<br>Home for<br>Thanksgiving | Overall |
| --- | --- | --- | --- | --- |
|  | n = 3,652 | n = 2,063 | n = 2,055 | n = 7,905 |
| **Median age in years, (IQR)** | 50 (35, 66) | 42 (29, 60) | 40 (30, 57) | 45 (32, 63) |
| **Male gender, n (%)** | 1,824 (50.2) | 1,035 (50.5) | 1,047 (51.2) | 3,965 (50.5) |
| **Race/Ethnicity, n (%)** |  |  |  |  |
| *White* | 1,948 (53.7) | 1,032 (50.3) | 1,182 (57.7) | 4,207 (53.6) |
| *Black/African American* | 506 (13.9) | 293 (14.3) | 215 (10.5) | 1,033 (13.2) |
| *Hispanic/Latino* | 781 (21.5) | 582 (28.4) | 542 (26.4) | 1,952 (24.9) |
| *Asian/Pacific Islander* | 326 (9.0) | 110 (5.4) | 82 (4.0) | 523 (6.7) |
| **Annual household income, n (%)** |  |  |  |  |
| < \$20,000 | 504 (14.8) | 277 (14.2) | 229 (11.6) | 1,036 (14.0) |
| \$20,000 – 39,000 | 614 (18.0) | 348 (17.9) | 354 (18.0) | 1,334 (18.0) |
| \$40,000 – \$49,000 | 308 (9.1) | 169 (8.7) | 196 (10.0) | 680 (9.2) |
| \$50,000 – \$69,000 | 622 (18.3) | 335 (17.2) | 373 (19.0) | 1,345 (18.1) |
| ≥ \$70,000 | 1,355 (39.8) | 819 (42.1) | 815 (41.4) | 3,018 (40.7) |
| **Mode of Thanksgiving travel** |  |  |  |  |
| *Car* |  |  | 1,750 (90.5) |  |
| *RV* |  |  | 28 (1.5) |  |
| *Train* | N/A | N/A | 42 (2.2) | N/A |
| *Bus* |  |  | 30 (1.6) |  |
| *Plane* |  |  | 72 (3.7) |  |
| *Boat* |  |  | 11 (0.5) |  |
| **Median size of Thanksgiving dinner, n (IQR)** | 1 (1, 3) | 4 (3, 6) | 5 (3, 8) | 3 (1, 5) |
| **COVID-19 symptoms in prior 2 weeks, n (%)** | 235 (6.5) | 178 (8.7) | 344 (16.9) | 774 (9.9) |
| **Tested for SARS-CoV-2 in prior 2 weeks, n (%)** | 313 (8.6) | 227 (11.1) | 470 (22.9) | 1,032 (13.2) |
| **Tested positive for SARS-CoV-2 in prior 2 weeks, n (%)\*** | 41 (13.1) | 47 (20.7) | 185 (39.4) | 278 (27.1) |
| **Median number of non-essential activities in prior 2 weeks, (IQR)** | 1 (0, 2) | 3 (1, 6) | 4 (2, 11) | 2 (0, 5) |
| **Masking when visiting with friends and family n (%)\*\*** |  |  |  |  |
| *Never* | 289 (27.8) | 410 (33.9) | 595 (37.2) | 1,305 (33.4) |
| *Sometimes* | 536 (51.5) | 636 (52.6) | 827 (51.8) | 2,035 (52.2) |
| *Always* | 215 (20.7) | 163 (13.5) | 176 (11.0) | 562 (14.4) |

\*among persons who received a SARS-CoV-2 PCR test in the prior 2 weeks
\*\*among 3,889 persons who reported visiting with friends or family either outdoors or indoors in the prior 2 weeks
Note: Numbers may not sum to total if there were participants who elected not to answer a given question.

## Data Availability

The data that support the findings of this study are available from the corresponding author upon reasonable request.

